# Modelling the Longitudinal Dynamics of Paranoia in Psychosis: A Temporal Network Analysis Over 20 Years

**DOI:** 10.1101/2023.01.06.23284268

**Authors:** J.M. Barnby, J.M.B. Haslbeck, R. Sharma, C. Rosen, M. Harrow

## Abstract

Paranoia is a highly debilitating, core element of psychosis, although is poorly managed. Theories of paranoia mostly interface with short-scale or cross-sectional data models, leaving the longitudinal course of paranoia underspecified. Here, we develop an empirical characterisation of two aspects of paranoia - persecutory and referential delusions - in individuals with psychosis over 20 years. We examine delusional dynamics by applying a Graphical Vector Autoregression Model to data collected from the Chicago Follow-up Study (n=135 with a range of psychosis-spectrum diagnoses). We adjusted for age, sex, IQ, and antipsychotic use. We found that referential and persecutory delusions are central themes, supported by other primary delusions, and are strongly autoregressive – the presence of referential and persecutory delusions is predictive of their future occurrence. In a second analysis we demonstrate that social factors influence the severity of referential, but not persecutory, delusions. We suggest that persecutory delusions represent central, resistant states in the cognitive landscape, whereas referential beliefs are more flexible, offering an important window of opportunity for intervention. Our data models can be collated with prior biological, computational, and social work to contribute toward a more complete theory of paranoia and provide more time-dependent evidence for optimal treatment targets.

**Graphical Abstract:** 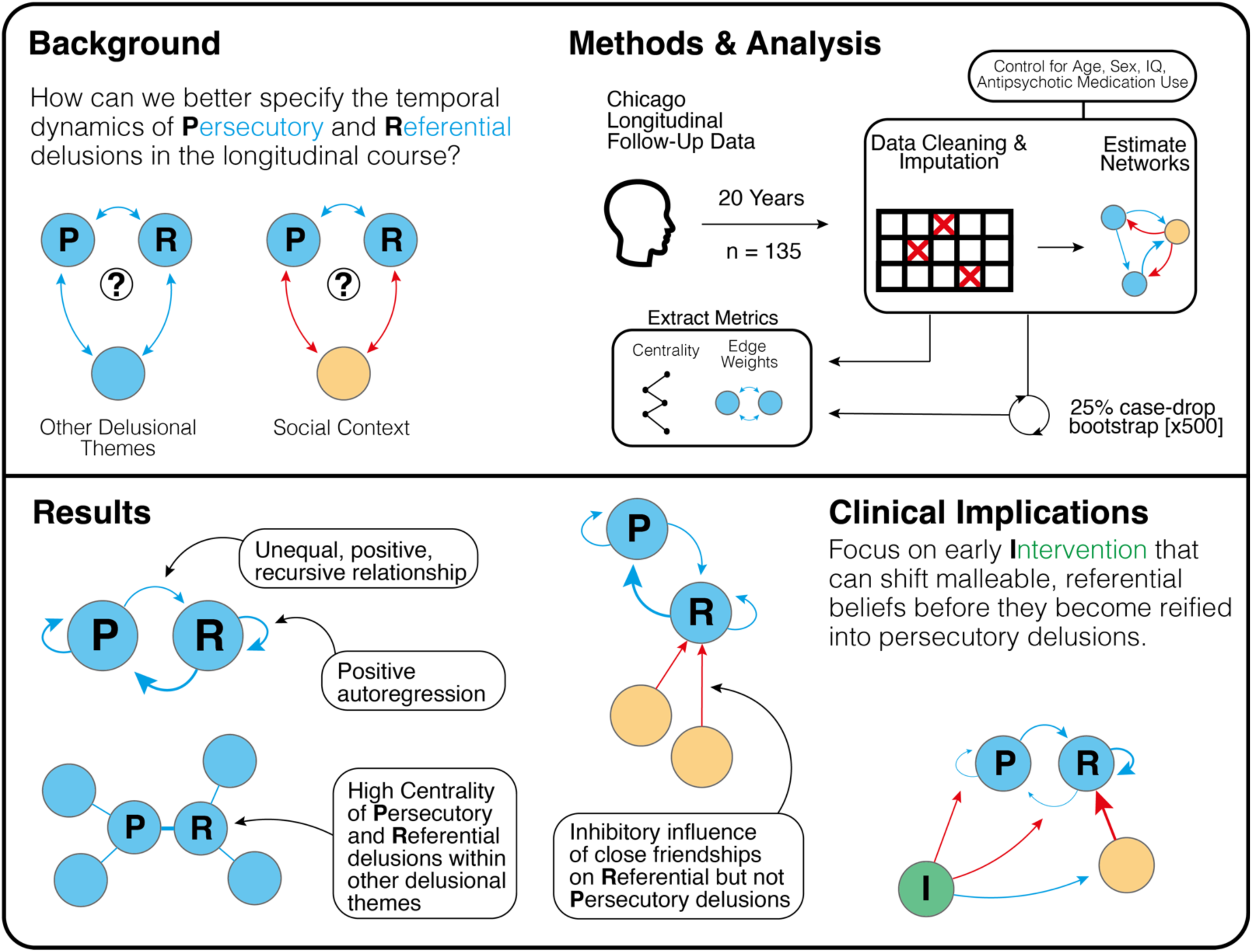

**Highlights:**

- Persecutory and referential delusions are central themes amongst primary delusions in chronic psychosis.
- Persecutory and referential delusions share a recursive relationship and are both strongly and positively autoregressive.
- Greater number and quality of friends reduce referential, but not persecutory, delusions.
- Our formal data model can be used as a test bed and framework for clinical intervention.

## 1.0 Introduction

Delusions cause profound physical and social disability, but current treatments are mostly unsuccessful. Delusions are transdiagnostic, noted to occur in around a third of those admitted to psychiatric wards (Jorgensen & Jensen, 1994), increasing the risk of poverty, unemployment, loneliness, homelessness and incarceration (Morgan et al., 2008). While approximately 70%-85% of individuals cease to be delusional after a year (Appelbaum et al., 2004; Gupta et al., 1997; Morgan et al., 2014), only a third of are expected to demonstrate good clinical response to current treatment (Morrison et al., 2020). In the likelihood that treatment is unsuccessful, delusions may remain over three (Dollfus & Petit, 1995), seven (Harrow et al., 1995), ten (Morgan et al., 2014), and twenty years (Harrow & Jobe, 2010; Harrow et al., 2012; Rosen et al., 2022). Common approaches to characterising delusional themes within psychosis rely on the prevalence of independent delusional themes (e.g. Collin et al., 2023; Harrow & Jobe, 2008), without examining their temporal causal structure.

Developing cost-effective and beneficial treatments for delusions requires strong formal theories that can identify windows of opportunity for intervention. Formal theories provide tools to improve knowledge (Robinaugh et al., 2021) and replicability (Borsboom et al., 2021a; Oberauer & Lewandowsky, 2019), allow transparency for falsification (Guest & Martin, 2020), and in clinical settings aspire to create scaffolds to guide personalised treatment (Huys et al., 2016). For example, computational models have been useful to formally characterise learning changes in mood and anxiety disorders (Pike & Robinson, 2020) and explain changes to social interaction in psychopathology (Barnby et al., 2023). Likewise, Artificial Neural Networks have proved useful as a model of aberrant perception (Keshavan & Sudarshan, 2017). Another popular framework, and one that addresses the complex, time and context dependent nature of psychopathology (Fried & Robinaugh, 2020; Fried, 2022), are complex system approaches. These suggest that psychopathology arises from dysfunctional interactions between mental states over time, which are causally connected through biological, cognitive and social mechanisms (Borsboom, 2008; 2017; 2021b).

Particularly, there has been a recent effort to formalise the temporal dynamics of paranoia to better identify targets for preventative clinical support. Paranoia is the most common delusional theme (Brakoulias & Starcevic, 2008; Cannon & Kramer, 2012; Collin et al., 2023; Paolini et al., 2016; Picardi et al., 2018), and increases social withdrawal, suicide attempts, and suicidal ideation, and reduces happiness, physical health, and social functioning (Freeman et al., 2011). Paranoia is structured hierarchically and along a continuum, with common referential beliefs (e.g. being talked about behind one’s back) being subordinate to rarer and more intense persecutory beliefs about intentional threat (e.g. believing that harm is occurring or will occur; Bebbington et al., 2013; Freeman et al., 2014; Freeman, 2016; Freeman et al., 2021). Referential beliefs may be especially sensitive to interpersonal dynamics (Bell & O’Driscoll et al., 2018; Hajdúk et al., 2019; Moffa et al., 2017); as paranoia gets more severe, beliefs may become untethered from the social environment (Raihani & Bell, 2019). Descriptive theory highlights the importance of genetic risk, and neurodevelopmental and social adversity in the production of severe paranoia (Murray & Howes, 2014).

Models of paranoia’s temporal nature currently focus on short-scale and cross-sectional observations, but its longitudinal course is largely unknown. Lag-models of subclinical paranoia over short time scales suggest it is autoregressive (paranoia in the moment predicts paranoia later on; Kuipers et al., 2018) and is sensitive to social stress, bullying, loneliness, and closeness to others (Contreras et al., 2020; 2022; Hermans et al., 2020), increasing the risk of more severe threat beliefs (Myin-Germeys et al., 2001; Veling et al., 2016). In clinical populations, paranoia is temporally predicted by sleep disturbances (Kasanova et al., 2020), affective fluctuations (Thewissen et al., 2011), and stress sensitivity (Reninghaus et al., 2016). Despite some exceptions (e.g. Bird et al., 2017; Fowler et al, 2012; Vorontsova et al., 2013), developing prospective, sophisticated data models to inform theory has been challenging since cohort data are difficult to collect.

A popular method to understand the temporal interrelationship of psychiatric phenomena, and one that interfaces with complex systems approaches to psychopathology, is network analysis. Network analytic data models empirically test, predict, and establish causal chains of target phenomena (Haslbeck et al., 2021), able to statistically implement the conditional relationships between nodes (symptoms) connected by edges (directed and undirected relationships) within a system (psychopathological phenomena). The resultant structures provide empirical weights around the centrality, stochasticity, and strength of symptoms within psychiatric syndromes (Epskamp, 2020; Marsman et al., 2018), allowing prediction, generation, and development of formal theory.

Here, we apply a complex systems approach implemented with network analysis to characterise the temporal evolution of two key components of severe paranoia, referential and persecutory delusions, over a 20-year period. This is with the overall aim to develop a more precise formal theory of the positive symptoms of psychosis. Using data collected from the Chicago Follow-up Study, we used lag-1 graphical vector autoregression models (GVAR; Epskamp et al., 2018) to examine interrelationships between paranoia and other primary delusions within and between individuals and characterise their centrality in chronic psychosis. To better understand the sensitivity of persecutory and referential delusions to social context we included social factors in a secondary analysis.

## 2.0 Methods

### 2.1 Participants and measures

This study reports findings examining persecutory delusions and delusions of reference, their associations with other delusions and social factors, in persons with psychosis over 20 years in the Chicago Follow-up Study (CFS). The CFS is a prospective, naturalistic, longitudinal research program designed to study psychopathology, neurocognition, and recovery in persons diagnosed with schizophrenia, other psychosis and non-psychotic depression (Harrow et al., 2005; Jobe and Harrow, 2005; Harrow et al., 2008; Strauss et al., 2010; Rosen et al., 2011; Lopez-Silva et al., 2022). The study was approved by the University of Illinois at Chicago Institutional Review Board (IRB#1997–0053). All participants signed informed consent prior to the initiation of study procedures and at each subsequent follow-up. The sample consisted of 555 participants who were recruited at index hospitalization and studied over six follow-ups at approximately 2, 4.5, 7.5, 10, 15, and 20 years later. We focus on 135 participants who had data recorded for our variables of interest at four or more follow up time point (see Supplementary Materials for missingness analysis).

Diagnosis was derived using the Diagnostic and Statistical Manual Version III criteria (American Psychiatric Association, 1980) and structured clinical interviews such as the Schedule for Affective Disorders and Schizophrenia (SADS; Endicott and Spitzer, 1978) and the Schizophrenia State Inventory (Grinker Sr and Harrow, 1987) and collateral information from hospital medical records, clinical staff and family members when available. Primary delusions (Persecutory, Reference, Bizarre, Grandiose, Religious, Thought Dissemination) were evaluated at each follow-up over 20-years using the SADS interview (SADS; Endicott and Spitzer, 1978). The SADS is a semi-structured diagnostic interview administered by trained clinicians and captures a detailed description of psychosis-relevant features over the past month prior to follow-up assessment using DSM relevant psychopathological dimensions. Delusions were classed as (1) absent, (2) equivocal (weak or occurring infrequently), or (3) definitely present. The Harrow Functioning Questionnaire was also administered at each follow-up and included the evaluation of social and work functioning through a structured interview (Grinker and Harrow, 1987; Harrow et al., 1997; Racenstein et al., 1999) and administered by trained research assistants who were blind to the diagnosis of participants. The evaluation of social functioning included items Total Social Satisfaction, Quality of Friends, Time with Friends, and Number of Close Friends at each of the six follow-up time points. For these items, participants were rated from 1 to 5, with 1 indicating good function and 5 indicating bad function. For analysis these scores were reversed, so that 5 represented good, and 1 bad. All follow-up measures were administered by trained raters in a clinical setting who were blinded to previous ratings and diagnoses (see Harrow et al., 2021; Rosen et al., 2022 for a detailed description of rating and follow-up schedules).

### 2.2 Lag-1 Panel Graphical Vector Auto-Regressive (GVAR) Analysis

To quantify the temporal (within individuals), between-individual, and relationships between residuals (contemporaneous: relationships between nodes averaged over time and averaged across the sample) of delusions and their relationships with social factors, we used Lag-1 Panel Graphical Vector Auto-Regressive (GVAR) analyses (Epskamp, 2020), estimated with the package ‘psychonetrics’ (Epskamp, 2021, v 0.10) in R (V 4.0.0) using the function ‘*panelgvar’* for estimation of panel data.

In these network models, nodes represent variables and edges represent their relationship when conditioned on all other nodes in the network in a number of fixed measurement occasions. In the case of between-subject and contemporaneous networks, these are undirected relationships. In the case of temporal networks, edges are directed (e.g., node *A -> B*) and conditioned upon all nodes at time *t*, as well as all nodes at *t-1* including itself (Epskamp, 2020). This uses a model drawing upon fixed effect lag-*k* variance-covariance matrices estimated from the data. Edges within temporal networks satisfy the condition that cause precedes effect, which is indicative of causality (Granger, 1969). For example, if a node, *i,* positively predicted itself (it is autoregressive) it means that scores on *i* at *t-1* predicts elevated scores at *i* at time *t.* If node *i* positively predicted another node, *j*, it means that scores on *i* at *t-1* predicts elevated scores on *j* at time *t*.

We estimated two different models: a first which only included the delusion variables, and a second which included the delusion variables and social variables. In all networks we utilised the unweighted least-squares (ULS) estimator in the ‘psychonetrics’ package due to our smaller sample and asymmetric data conditions at each time point (Li, 2016). For all missing data we imputed values using linear interpolation implemented in the ‘imputeTS’ package (Tomas et al., 2015). To control for confounders, at each time point (*t*) we regressed out age (*Age^t^*), whether a participant was receiving antipsychotic medication (*Medication^t^*), sex (*Sex^t^*^=0^) at baseline, and IQ (*IQ*^*t*=0^; see Eq. 1):

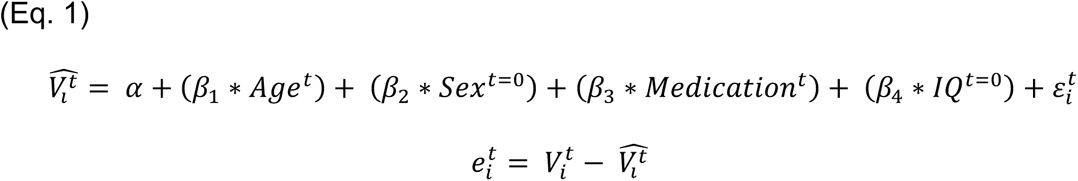

The resulting residuals (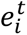) from each equation (observed values, 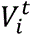, minus predicted values, 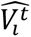, for each variable, *i,* at each time point) were then retained as nodes for the models. We then applied scaling (using the ‘*scale*’ function in R) to 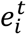 to ensure all nodes and edges were assessed on the same dimension and any mean trends removed. We then extracted the beta (temporal), omega-zeta-within (contemporaneous), and omega-zeta-between (between-individuals) subject matrices. For each of the networks we generated simulated data using the approximated full model structure and refitted the model on the simulated data to estimate model recovery. Residual variance was estimated using Cholesky decomposition. Model fit was assessed using the following fit statistics: we used relative fit indices (Tucker Lewis Index [TLI]) and the non-centrality-based indices (Comparative Fit Indices [CLI]) and Root Mean Square Error Approximation (RMSEA) with 95% confidence intervals (95%CI). Absolute fit indices are reported (Chi-Square). However, due to the metric’s sensitivity to sample size, Chi-square estimates were not interpreted. RMSEA < 0.05 indicate excellent fit; RMSEA < 0.10 indicate good fit; For TLI and CFI, values 0.90 – 0.95 are considered as accepted cut offs for good fit (Sivo et al., 2006).

To evaluate robustness of the estimates and to avoid overfitting in our networks, we computed the stability of edges within each full fitted network using bootstrapping procedures: over 500 iterations, 25% of the sample was randomly held out and the full model refitted on the remaining 75% of participants. Within each iteration the selected data imputed, each variable regressed against confounders, and scaled in the same manner as in the full model to control for errors and variance within the data cleaning and scaling process. The averaged edge weights and 95% confidence intervals (CI) over all 500 iterations were retained and reported. All edges that had 95%CI crossing zero were forced to 0.

### 2.3 Delusion network

We included all primary delusion variables at each of the six time follow up time-points into the network model (Persecutory - PER, Reference - REF, Bizarre - BIZ, Grandiose - GRD, Religious - REL, Thought Dissemination - THD) to examine their interrelationship in the chronic phase of disorder. Higher values indicate more severe delusions. Nihilism was originally included in our analysis plan although led to model fitting issues and so was rejected in favour of a leaner, better fitting network model which may have been more interpretable. See Supplementary Materials (Figure S2) for the full model fit with Nihilism included.

### 2.4 Social network

We included REF and PER, and additionally included Social Satisfaction (SAT), Quality of Friends (QOF), Time with Friends (TWF), and Number of Close Friends (NCF) at each of the six follow-up time points.

## 3.0 Results

Our cohort (n = 135) was followed up over 20 years at 6 follow ups after initial contact with psychiatric services at index hospitalization (see Figure 1). Primary delusions were assessed using the Schedule for Affective Disorders and Schizophrenia (SADS), and social function was assessed with the Harrow Functioning Questionnaire (HFQ).

**Figure 1.**
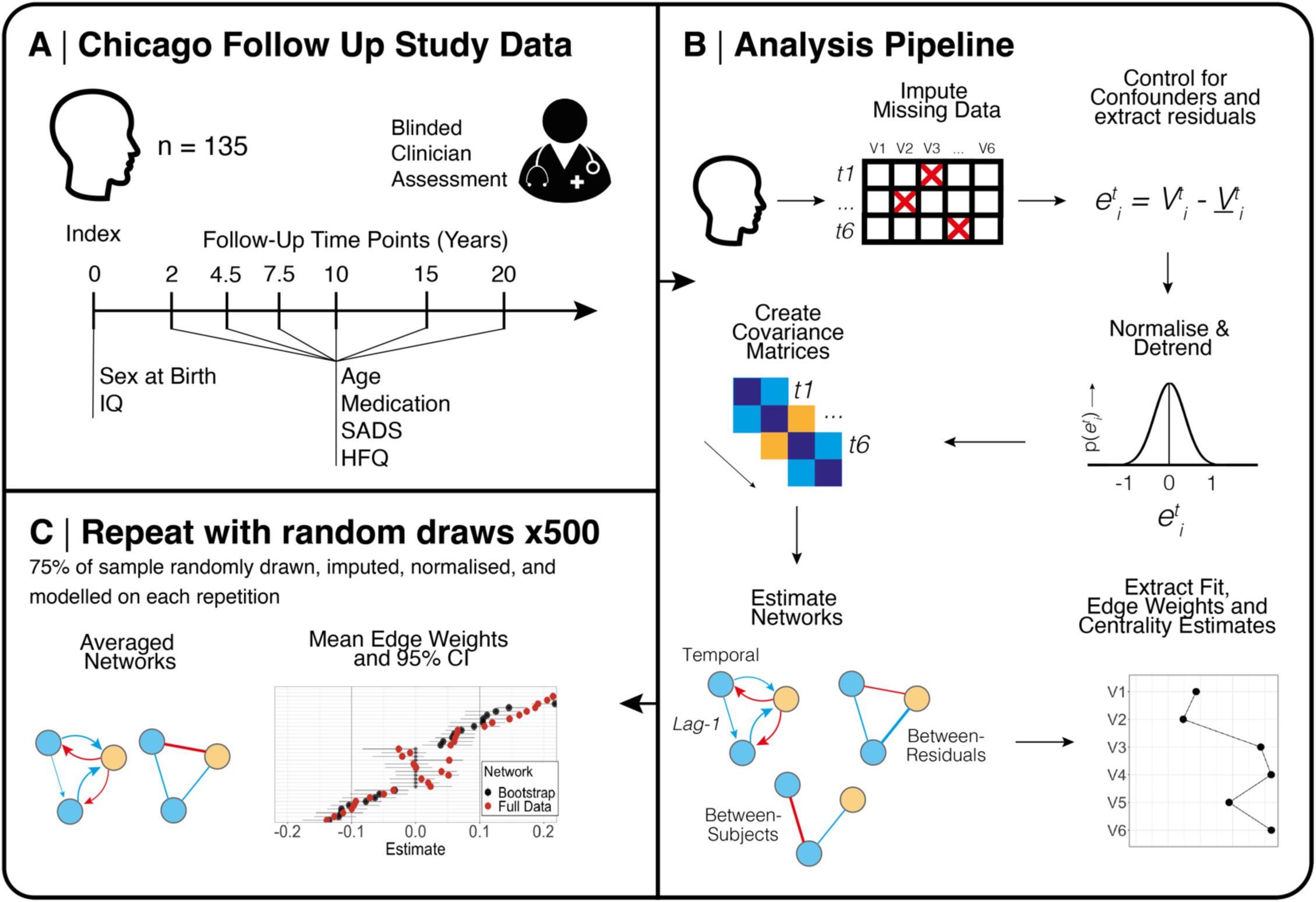
Summary of study design and analysis pipeline. Random draws consisted of randomly sampling 75% of the full sample and estimating primary and secondary networks. Each random draw performed independent imputation, control analysis, residual extraction, normalisation, and modelling. All average edge weights were forced to zero if 95% confidence intervals crossed zero. SADS = Schedule for Affective Disorders and Schizophrenia. HFQ = Harrow Functioning Questionnaire.

52.5% were male and had on average 13.8 years of education (SD = 3.3). Participants were mostly in their mid-twenties at the first 2-year follow up (mean = 25.5, SD = 4.8). 57.8% of the cohort were not on antipsychotic medication at the 2-year follow-up, and over time, antipsychotic medication was reduced across the sample. See Table 1 for a summary of demographic variables, primary delusion prevalence and social factors. See Figure S1 and S2 for raw and normalised between-subject change in delusions over time.

**Table 1.**
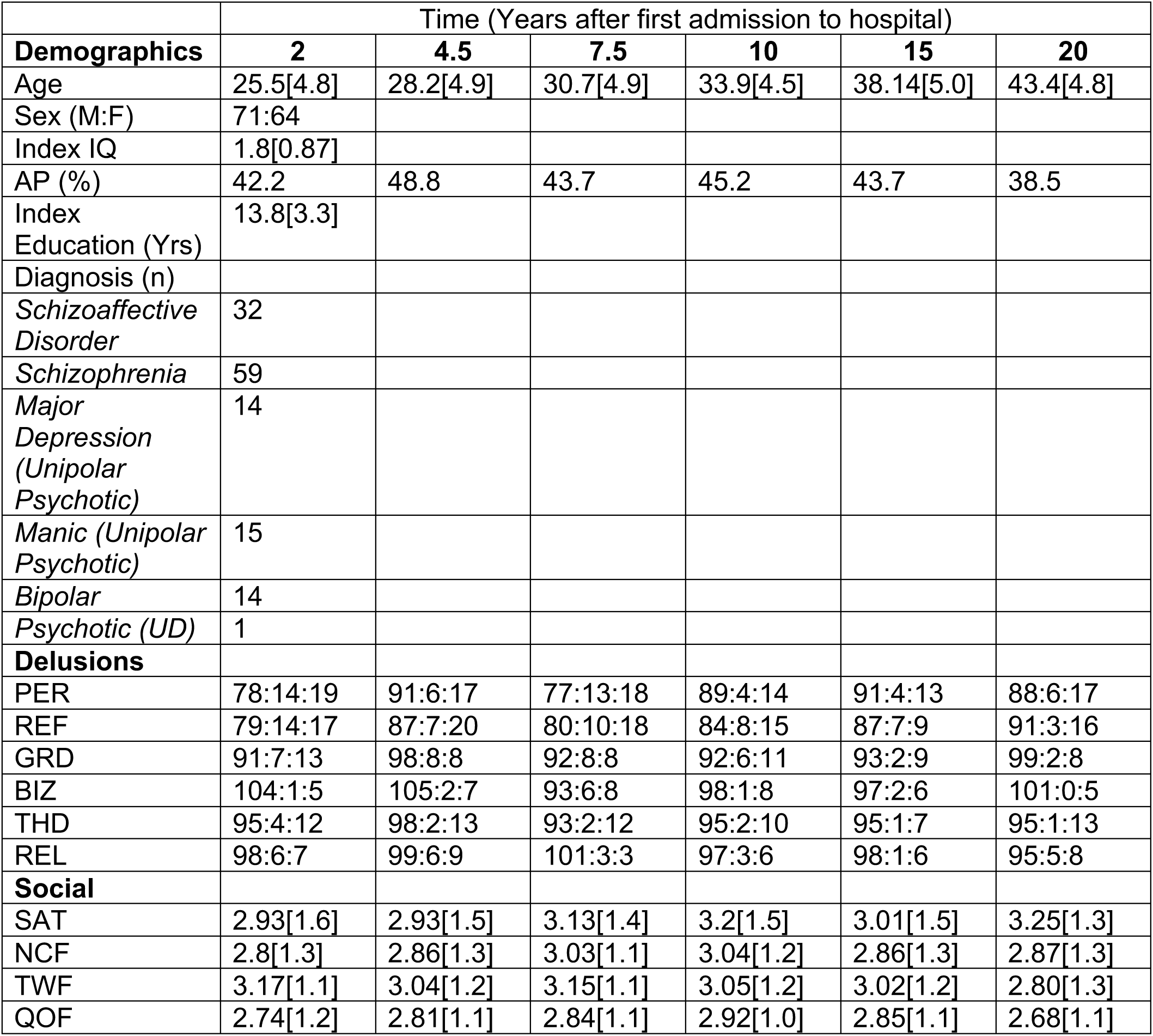
Summary statistics of the total sample. N = 135 excluding missing cases at each time point and/or for each variable (total missingness = 16-17%; see Supplementary Material). AP = Antipsychotic medication. IQ was measured as 1 = Severe Impairment, 2 = Mild Impairment, 3 = IQ within normal range. Delusions were measured using an ordinal scale prior to being regressed against confounding variables: 1 = not present, 2 = borderline, 3 = present; they are presented on the table as the number of participants occupying each classification, i.e. n_1_:n_2_:n_3_. Social variables, Age, IQ, and Education are all reported as means and standard deviations [sd]. THD = Thought dissemination, REF = Reference, PER = Persecutory, BIZ = Bizarre, GRD = Grandiose, REL = Religious, NCF = Number of close friends, QOF = Quality of friends, TWF = Time with friends, SAT = Social satisfaction, UD = Undifferentiated.

### 3.1 Primary Analysis (Delusions-Only Network)

Primary delusions input into the model included Persecutory (PER), Referential (REF), Religious (REL), Grandiose (GRD), and Bizarre Delusions (BIZ), and Thought Dissemination (THD). A saturated model had better fit than a sparse model (*X*^2^(58) = 2607.30, p < .001); the former being a densely connected network with all available edges, and the latter being a network with pruned edges. The saturated network demonstrated good fit (RMSEA = .072 [95%CI: .064, .079]; *χ*^2^(618) = 1048.54, p < .001; CFI = .90; TLI = .90). Simulated data (n = 135) using the estimated full delusion model structure was generated and refitted to estimate recoverability, demonstrating excellent fit (RMSEA = 0.048 [95%CI: 0.044, 0.051]; *X*^2^(618) = 1326.38, p < .001; CFI = .96; TLI = .96). We estimated a temporal and residual network from the data (Figure 1; Table 2A) and performed a 25% case-dropping bootstrap (Table S1 & Figure 2D) to assess the stability of approximated edges. All edges showed high stability.

**Figure 2.**
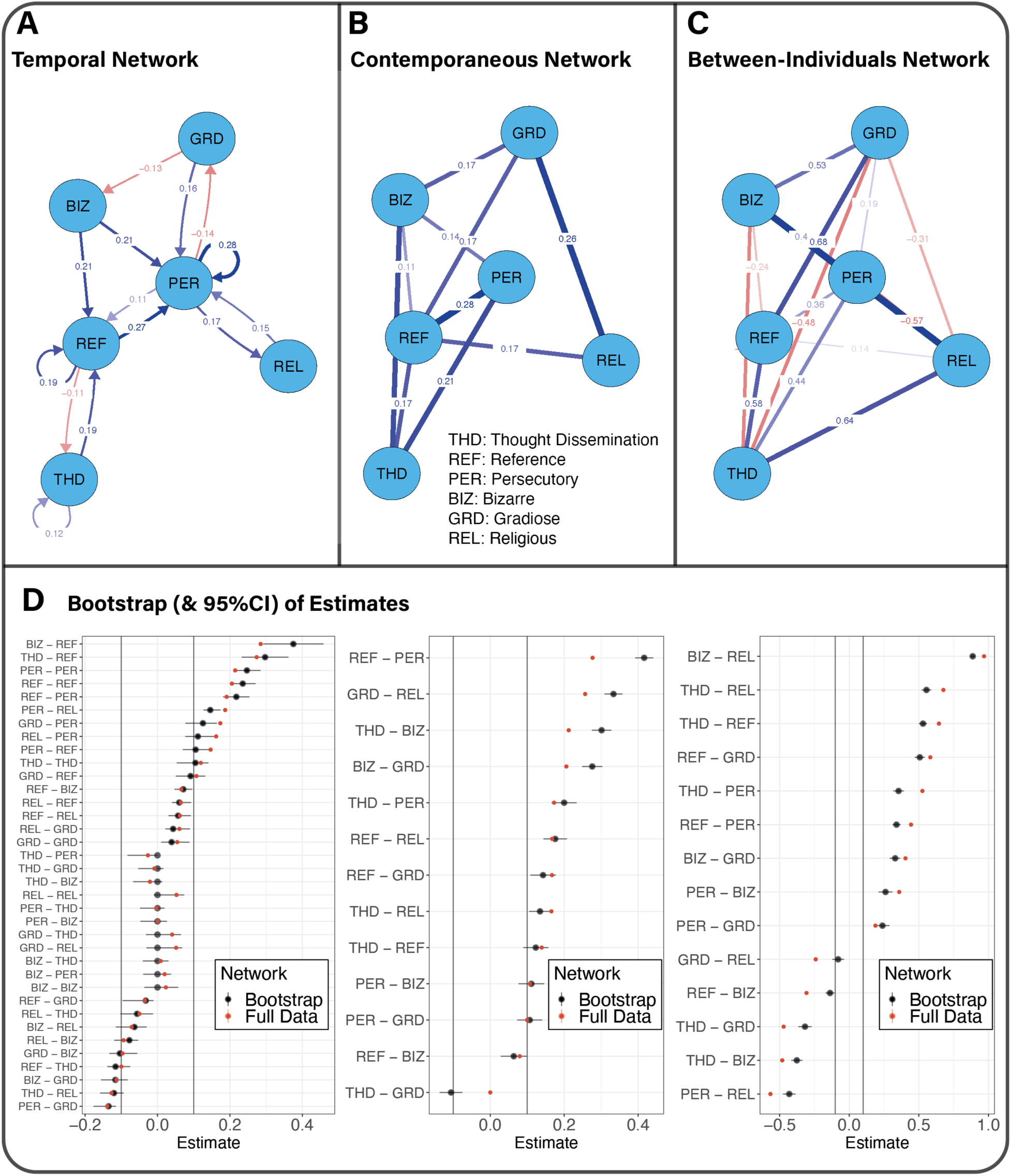
Full data estimation and bootstrapping for the delusions-only network. For Panels A-C, a threshold with an absolute value of 0.1 was applied to remove all small edges from the visualization. (A) Temporal relationships between nodes. (B) Contemporaneous relationships between nodes. (C) Between-individual relationships between nodes. (D) Estimates of full data relationships (n=135; red) and bootstrapped estimates (500 repetitions; black) with 95% confidence intervals. Edges with 95%CI crossing zero were forced to zero. For each bootstrap, 75% of the sample was randomly drawn without replacement and missing data imputed before being modelled in the same manner as the full data approximations.

**Table 2.**
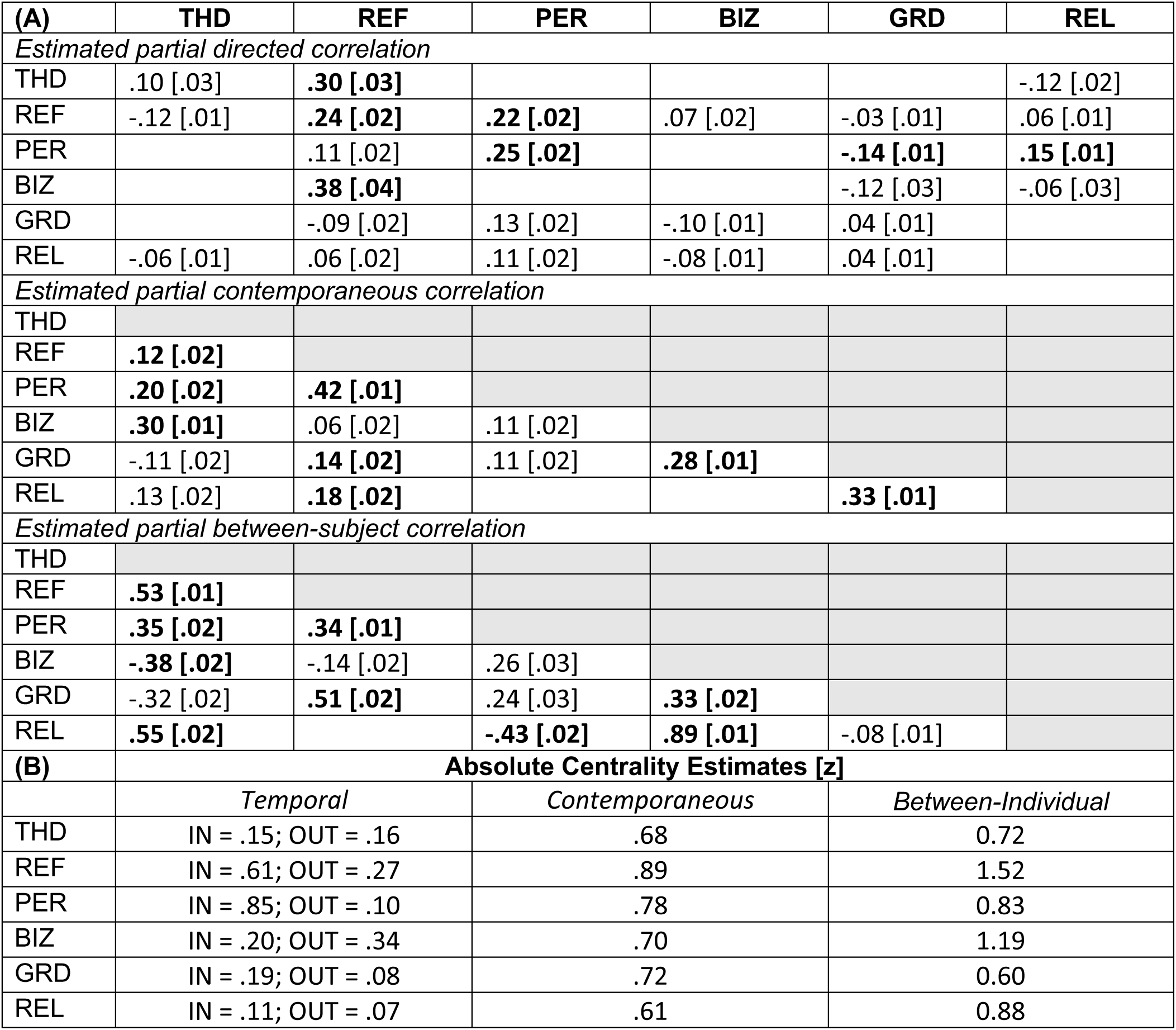
Numeric bootstrapped results of the *GVAR* analysis of the delusions-only data. (A) All estimates are partial correlation coefficients from bootstrapping which have 95%CI intervals that do not cross zero. Standard error of the mean is recorded within square brackets. **Bold** = edges are also significant at least at the p<0.05 level in the model fitted on all (n=135) data. All edges in the full (n=135) data survived case-dropped bootstrapping following 500 repetitions, aside from the unidirectional relationship from BIZ to PER in the full data. THD = Thought dissemination, REF = Reference, PER = Persecutory, BIZ = Bizarre, GRD = Grandiose, REL = Religious. (B) Centrality estimates of each full network model. Numbers indicate the total sum of weights either to (IN) or from (OUT) of nodes. For case-dropping bootstrap centrality estimates see Table S1.

| (A) | THD | REF | PER | BIZ | GRD | REL |
| --- | --- | --- | --- | --- | --- | --- |
| *Estimated partial directed correlation* |  |  |  |  |  |  |
| THD | .10 [.03] | **.30 [.03]** |  |  |  | -.12 [.02] |
| REF | -.12 [.01] | **.24 [.02]** | **.22 [.02]** | .07 [.02] | -.03 [.01] | .06 [.01] |
| PER |  | .11 [.02] | **.25 [.02]** |  | **-.14 [.01]** | **.15 [.01]** |
| BIZ |  | **.38 [.04]** |  |  | -.12 [.03] | -.06 [.03] |
| GRD |  | -.09 [.02] | .13 [.02] | -.10 [.01] | .04 [.01] |  |
| REL | -.06 [.01] | .06 [.02] | .11 [.02] | -.08 [.01] | .04 [.01] |  |
| *Estimated partial contemporaneous correlation* |  |  |  |  |  |  |
| THD |  |  |  |  |  |  |
| REF | **.12 [.02]** |  |  |  |  |  |
| PER | **.20 [.02]** | **.42 [.01]** |  |  |  |  |
| BIZ | **.30 [.01]** | .06 [.02] | .11 [.02] |  |  |  |
| GRD | -.11 [.02] | **.14 [.02]** | .11 [.02] | **.28 [.01]** |  |  |
| REL | .13 [.02] | **.18 [.02]** |  |  | **.33 [.01]** |  |
| *Estimated partial between-subject correlation* |  |  |  |  |  |  |
| THD |  |  |  |  |  |  |
| REF | **.53 [.01]** |  |  |  |  |  |
| PER | **.35 [.02]** | **.34 [.01]** |  |  |  |  |
| BIZ | **-.38 [.02]** | -.14 [.02] | .26 [.03] |  |  |  |
| GRD | -.32 [.02] | **.51 [.02]** | .24 [.03] | **.33 [.02]** |  |  |
| REL | **.55 [.02]** |  | **-.43 [.02]** | **.89 [.01]** | -.08 [.01] |  |
| (B) | **Absolute Centrality Estimates [z]** |  |  |  |  |  |
|  | *Temporal* | *Contemporaneous* |  | *Between-Individual* |  |  |
| THD | IN = .15; OUT = .16 | .68 |  | 0.72 |  |  |
| REF | IN = .61; OUT = .27 | .89 |  | 1.52 |  |  |
| PER | IN = .85; OUT = .10 | .78 |  | 0.83 |  |  |
| BIZ | IN = .20; OUT = .34 | .70 |  | 1.19 |  |  |
| GRD | IN = .19; OUT = .08 | .72 |  | 0.60 |  |  |
| REL | IN = .11; OUT = .07 | .61 |  | 0.88 |  |  |

The strongest autocorrelations were associated with persecutory delusions (.28) and delusions of reference (.19, Table 2A; Figure 2A). Delusions of reference were observed to be a strong predictor of persecutory delusions at the next time point (.27), and less evidence was observed for the opposite direction (.11). The next most prominent unidirectional relationship was bizarre delusions and delusions of reference (.21) and delusions of persecution (.21). With respect to recursive edges, where relationships existed in both directions between two nodes, grandiose delusions were positively associated with persecutory delusions at the next time point (.16), and in contrast, persecutory delusions were negatively associated with grandiose delusions (-.14). Considering the centrality of items (total sum of weights to and from nodes), delusions of reference (z-score [z] = .61) and persecutory delusions (z = .85) had the strongest In-Expected influence, whereas bizarre delusions (z = .34) had the strongest Out-Expected influence (Table 2B).

Inspection associations between residuals, i.e., the partial correlation between nodes after conditioning on time and between-subject effects (Table 2A; Figure 2B), delusions of reference and persecutory delusions had the strongest positive relationship, followed by grandiose and religious delusions, and bizarre and persecutory delusions. Within this network, the most central items were delusions of reference (z = .89) and persecutory delusions (z = .78), followed by grandiose delusions (z = .72).

Between-individuals estimates (Table 2A; Figure 2C) suggested that bizarre and religious delusions shared the strongest positive relationships, followed by delusions of reference and persecutory delusions, and thought dissemination and religious delusions. Persecutory delusions and religious delusions shared the strongest negative association, followed by thought dissemination and grandiose delusions, and thought dissemination and bizarre delusions. The most central items were delusions of reference (z = 1.52) and bizarre delusions (z = 1.19), followed by religious (z = .88) and persecutory (z = .83) delusions.

3.2 Secondary Analysis (Social Network)

Variables input into the secondary analysis were Number of Close Friends (NCF), Quality of Friends (QOF), Time with Friends (TWF), Social Satisfaction (SAT) as well as PER and REF delusions. A saturated model had better fit than a sparse model (*X*^2^(59) = 1008.71, p < .001). The saturated network had excellent fit (RMSEA ∼ 0; *X*^2^(630) = 556.58, p = .96; CFI = 1; TLI = 1.02). Simulated data using the estimated full delusion model structure was generated and refitted to determine recoverability, demonstrating excellent fit (RMSEA ∼ 0; *X*^2^(1670) = 1562.44, p = 1; CFI = 1.00; TLI = 1.01). We estimated a temporal, contemporaneous, and between-subject network from the data (Figure 3; Table 3) and performed a 25% case-dropping bootstrap to assess the stability of approximated edges. All edges displayed excellent stability.

**Figure 3.**
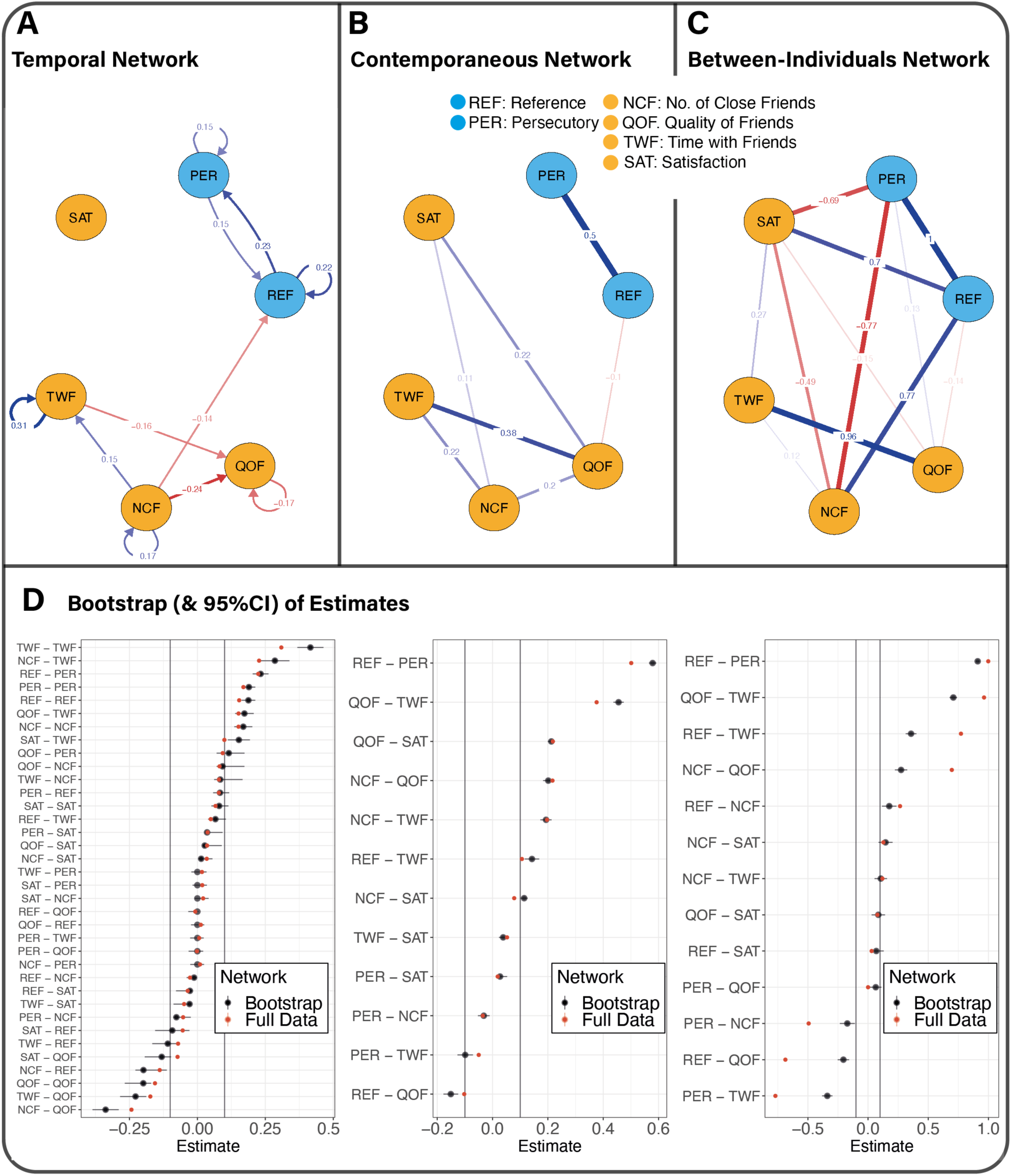
Full data estimation and bootstrapping for the social network. (A) Temporal relationship between nodes. (B) Contemporaneous relationship between nodes. (C) Between-individual relationships between nodes. (D) Estimates of full data relationships (n=135; red) and bootstrapped estimates (500 repetitions; black) with 95% confidence intervals. For each bootstrap, 75% of the sample was randomly drawn without replacement and missing data imputed before being modelled in the same manner as the full data approximations. For Panels A-C, a threshold of 0.1 was applied to remove all small edges.

**Table 3.** Numeric bootstrapped results of the *GVAR* analysis of the social & delusions data. (A) All estimates are partial correlation coefficients from bootstrapping which have 95%CI intervals that do not cross zero. Standard error of the mean is recorded within square brackets. **Bold** = edges were significant at least at the p<0.05 level in the model fitted on all (n = 135) data. All edges in the full data (n = 135) survived case-dropping bootstrapping after 500 repetitions. REF = Reference, PER = Persecutory, NCF = Number of close friends, QOF = Quality of friends, TWF = Time with friends, SAT = Social satisfaction. (B) Centrality estimates of each full network model. Numbers indicate the total sum of weights either to (IN) or from (OUT) of nodes. For case-dropping bootstrap centrality estimates see Table S1.

| (A) | REF | PER | NCF | QOF | TWF | SAT |
| --- | --- | --- | --- | --- | --- | --- |
| *Estimated partial directed correlation* |  |  |  |  |  |  |
| REF | **.19 [.01]** | **.23 [.01]** | -.01 [.01] |  | .07 [.01] | -.03 [.00] |
| PER | **.08 [.01]** | .19 [.01] | -.08 [.01] |  |  | .04 [.00] |
| NCF | **-.20 [.04]** |  | **.17 [.02]** | **-.34 [.02]** | **.29 [.03]** | .01 [.01] |
| QOF |  | .12 [.03] | **.09 [.01]** | **-.20 [.01]** | .17 [.02] | .03 [.00] |
| TWF | -.11 [.04] |  | .08 [.02] | -.23 [.02] | **.42 [.02]** | -.03 [.01] |
| SAT | -.09 [.03] |  |  | -.13 [.02] | .15 [.02] | .08 [.00] |
| *Estimated partial contemporaneous correlation* |  |  |  |  |  |  |
| REF |  |  |  |  |  |  |
| PER | **.58 [.01]** |  |  |  |  |  |
| NCF |  | -.03 [.01] |  |  |  |  |
| QOF | **-.15 [.01]** |  | **.20 [.01]** |  |  |  |
| TWF | .14 [.01] | -.10 [.01] | **.19 [.01]** | **.46 [.01]** |  |  |
| SAT |  | .03 [.01] | .11 [.01] | **.21 [.01]** | .04 [.01] |  |
| *Estimated partial between-subject correlation* |  |  |  |  |  |  |
| REF |  |  |  |  |  |  |
| PER | .91 [.01] |  |  |  |  |  |
| NCF | .18 [.03] | -.17 [.03] |  |  |  |  |
| QOF | -.20 [.02] | .06 [.02] | .27 [.03] |  |  |  |
| TWF | .36 [.02] | -.34 [.02] | .11 [.03] | .71 [.02] |  |  |
| SAT | .07 [.03] |  | .15 [.03] | .09 [.03] |  |  |
| (B) | **Absolute Centrality Estimates [z]** |  |  |  |  |  |
|  | *Temporal* | *Contemporaneous* |  | *Between-Individual* |  |  |
| REF | IN = .22; OUT = .29 | .40 |  | 2.60 |  |  |
| PER | IN = .23; OUT = .15 | .50 |  | 2.58 |  |  |
| NCF | IN = .00; OUT = .53 | .52 |  | 2.15 |  |  |
| QOF | IN = .40; OUT = .00 | .69 |  | 1.38 |  |  |
| TWF | IN = .16; OUT = .15 | .59 |  | 1.35 |  |  |
| SAT | IN = .00; OUT = .00 | .32 |  | 2.29 |  |  |

Similarly to the delusions-only network we found an unequal but recursive relationship between delusions of reference and persecutory delusions; referential delusions and persecutory delusions predicted each other at the next time point (Table 3A; Figure 3A). Time with friends (.31) and number of close friends (.17) was strongly and positively autoregressive, whereas quality of friends showed the opposite (negative autoregressive) pattern (-.17). Number of close friends predicted fewer delusions of reference in the future (-.14); as number of close friends increased at the current measurement instance, delusions of reference were fewer in the next. However, there was no influence of social factors on persecutory delusions. There was also some evidence for quality of friends reducing delusions of reference (Table 3A). This may suggest that in one direction, a positive feedback loop may be initiated, from improved friendship quality to fewer delusions of reference; the opposite however may lead to a negative feedback loop toward a negative outcome, where heightened delusions of reference reduce the perceived quality of friends. Considering the absolute centrality of items, quality of friends (z = .40), and referential delusions (z = .29) had the strongest In-Expected influence, whereas number of close friends (z = .53) and delusions of reference (z = .23) had the strongest Out-Expected influence.

Inspecting associations between residuals (Table 3A; Figure 3B), quality of friends was negatively associated with delusions of reference, although shared no relationship with persecutory delusions. Quality of friends and time with friends were strongly positively associated. Within this network, the most central items were quality of friends (z = .65), and time with friends (z = .70). Between-individual edges (Table 3A; Figure 3C) demonstrated a strong negative association between overall life satisfaction and persecutory delusions, and number of close friends. Number of close friends was also negatively associated with persecutory delusions, but positively associated with delusions of reference. The most central items were delusions of reference (z = 2.60) and persecutory delusions (z = 2.58).

## 4.0 Discussion

In order to empirically characterise paranoia in chronic disorder we performed a GVAR panel analysis on a cohort of participants with psychosis-spectrum disorders over 20-years, controlling for age, sex, IQ, and antipsychotic medication status. A number of findings stood out: 1) relationships between persecutory and referential delusions were unequal, but positive and bidirectional; persecutory delusions were more strongly predicted by the presence of referential delusions, 2) both persecutory and referential delusions were strongly and positively autoregressive over time, 3) other delusional themes had the greatest influence on referential and persecutory delusions, and 4) the number of close friends and quality of friends had different effects on persecutory and referential delusions, with referential delusions being reduced but not persecutory delusions. Importantly, despite 20 years of clinical support, some individuals continued to present with delusions, and this was centred on persecutory and referential themes. Together, our data, analytic pipeline, and model structure can be used to generate predictions and test the influence of biological, social, and psychological causes on the formation and dissolution of severe paranoia over short and long-time scales.

Our results are consistent with cognitive models of paranoia and extend them to encompass the longitudinal dynamics of psychotic disorders. Cognitive models suggest that through affective activation, cognitive biases, and avoidance of social contact, referential beliefs may develop into persecutory beliefs (Freeman, 2016). Temporal models (Contreras et al., 2020; 2022; Hermans et al., 2020) and experience momentary assessment (Kasanova et al., 2020; Reininghaus et al., 2016; Thewissen et al., 2011) have probed the predictive factors leading to persecutory beliefs, although their dynamics over long time periods was unclear. Our findings support and extend these data and models, indicating the persistence of paranoia, that delusions of reference are more sensitive to social context, and that social environments may have little effect once referential themes have crystallised into persecutory delusions. Our analysis supports the proposal that social factors play a core role in the development of delusions, rather than being a corollary of them (Badcock et al., 2020).

Our data and model allow clear predictions about the directionality, centrality, and predictive factors of paranoia over long periods of time which can abductively generate formal theory. Referential and persecutory delusions in our model showed high centrality, autoregressive traits, and strong interrelationships, reflecting their strong influence in chronic psychosis. This may be conceptualised as an increase in the depth and resilience of their attractor basins within psychological state space, creating a self-perpetuating cycle (Scheffer et al., 2022). Causal factors in the maintenance and development of this dramatic shift in the belief landscape remain unclear, although may be further investigated with more clarity using the data model we present. We now discuss avenues for future research to unpick the causal mechanisms that may together build a more complete theory of paranoia.

The psychological and biological mechanisms of paranoid states have been addressed with computational modelling. According to formal process models of paranoia (Adams et al., 2022; Barnby et al., 2020a; 2022) and general models of delusional decision making (Baker et al., 2019; Deserno et al., 2020; Diaconescu et al., 2020; Erdmann & Mathys, 2022; Nassar et al., 2019) higher prior precision over threat beliefs about others, greater uncertainty about others, and resistance of threat beliefs to changes in an other’s behaviour may ’trap’ beliefs within a self-fulfilling cycle. Others are viewed as being more harmful, and evidence to the contrary has little impact on these beliefs. Dopaminergic function is important to coordinate these components, being implicated in the precision (Adams et al., 2022), volatility (Reed et al., 2020), and magnitude (Barnby et al., 2020b) of paranoia. As a future goal, understanding the neural dynamics underlying decision making in environments relevant to persecutory delusions is critical for understanding how resilient states may be implemented in the cortex and expressed in cognition.

Abuse and neglect during critical periods of development, such as early life and adolescence, can have a significant impact on the development of close social ties in adulthood and increase the risk of developing paranoia. This is particularly relevant for humans, who have a more fractal social network structure, where close relationships rely heavily on the establishment of stable trust and low threat (Dunbar, 2020). Disruptions during critical developmental stages, such as peer difficulties (Bird et al., 2021) and parental abuse (Brown et al., 2021) contribute to the risk of developing paranoia. In adulthood, coupled with genetic vulnerability and neurodevelopmental stress (Howes & Murray, 2014; Freeman, 2016), traumatic disruptions to the development of close secure attachments are linked to clinically-relevant paranoia (Bloomfield et al., 2021; Cosgrave et al., 2021; Humphrey et al., 2021). Developing longitudinal cohorts of adolescents in conjunction with formal dynamic models will be crucial to examine the causal effect of abuse and neglect on lasting adult paranoia.

In line with prior work, we propose clinical practice should focus on early interventions that can shift malleable, referential beliefs before they become reified into persecutory delusions. This may be addressed through interventions that focus on improving the quality and number of close social relationships (Lamster et al., 2017; Lim et al., 2018; Michalska da Rocha et al., 2018; Sündermann et al., 2014). Emphasis given to reducing stigma and increasing social support for individuals experiencing persistent paranoia may increase the likelihood of avoiding social isolation (Colizzi et al., 2020; Freeman et al., 2007); this is not only one of the many important aspects to peer-led recovery groups for individuals experiencing voices and/or other extreme states including non-consensus reality (Romme & Escher, 2019), but may have an impact on the exacerbation of psychotic symptoms more broadly (Hoffman, 2007; Marschall et al., 2020). This can in turn lead to reduced loneliness and the abatement of psychotic symptoms (Lim et al., 2018; Michalska da Rocha et al., 2018). Early approaches to address loneliness and paranoia using virtual reality cognitive behavioural therapy (VR-CBT) and network approaches to evaluate intervention success find that VR-CBT was able to break the autoregression of suspiciousness and reduce the recursion between loneliness and suspiciousness (Geraets et al., 2020). This may be a longer-term clinical goal to prevent relapse once rigid persecutory delusions have been reduced. We hope our analysis will improve policy guidance into the most appropriate targets for therapeutic intervention and highlight the risk of not taking early action when beliefs may be most flexible.

We note several limitations of our approach and dataset. To utilise as much data as possible we imputed missing values and normalised residuals after controlling for confounds. This ensured the same scale for all variables and allowed for missing data. This might mean that some edges, while internally consistent, may fluctuate far more than approximated. Nevertheless, bootstrapping procedures aimed to take uncertainty of the imputation process into account on each iteration by re-imputing data within each bootstrap sample. Secondly, it is unclear whether clinician ratings are reliable; clinicians were blind to previous ratings but may contain variance into what each would consider a ‘clear’ delusion versus a borderline case. Finally, the GVAR model assumes that the interval between measurement instances is constant and that the dynamics from each pair of time points is the same. The first assumption is violated in our data, because there is some variation between intervals. This leads to the situation that the estimated parameters are actually a weighted mixture of effects at different time scales. The second assumption is likely violated, since the study stretched 20 years and it is unlikely that all considered processes are invariant across the lifespan. However, the complexity of the model we could fit was limited by the available data and we therefore were unable to model effects dependent on equal time intervals time. Future work that is able to incorporate random-effects between time points for each participant and include time-varying associations would significantly strengthen the conclusions made from our analyses.

## Data Availability

Covariance matrices to estimate networks and all analysis code are available on GitHub: https://github.com/josephmbarnby/ParanoiaLongitudinalNetworkAnalysis.

## Conflicts of Interest

None to declare

## Funding

This work was supported in part by USPHS Grants MH-26341 and MH-068688 from the National Institute of Mental Health, USA (MH) and a Grant from the Foundation for Excellence in Mental Health Care G5014 (MH). JMBH has been supported by the gravitation project ‘New Science of Mental Disorders’ (www.nsmd.eu), supported by the Dutch Research Council and the Dutch Ministry of Education, Culture and Science (NWO gravitation grant number 024.004.016).

## Acknowledgments

The authors would like to sincerely thank all the individuals who participated in the Chicago Longitudinal Study as their contributions over the 20 years made this research possible.

## Notes

### Competing Interest Statement

The authors have declared no competing interest.

### Author Declarations

Ethics Committee/IRB of University of Illinois at Chicago gave ethical approval for this work

### Summary of Updates

Revised in response to peer review (Psychiatry Research)

