## Supplementary Materials for "Modelling the Longitudinal Dynamics of Paranoia in Psychosis: A Temporal Network Analysis Over 20 Years"

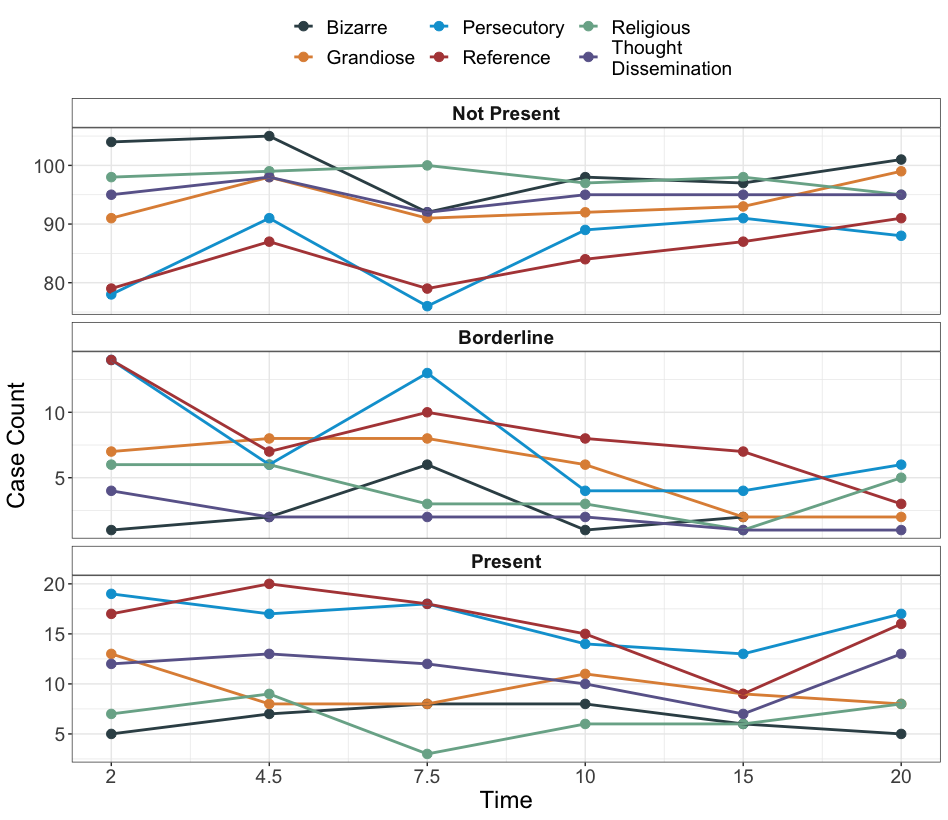

**Figure S1. Between-individual sum of cases for each timepoint.**

Participants were rated by a clinician as to whether delusions were not present, were borderline, or were clearly present. This data is not controlled for by age, sex, IQ, or medication status.

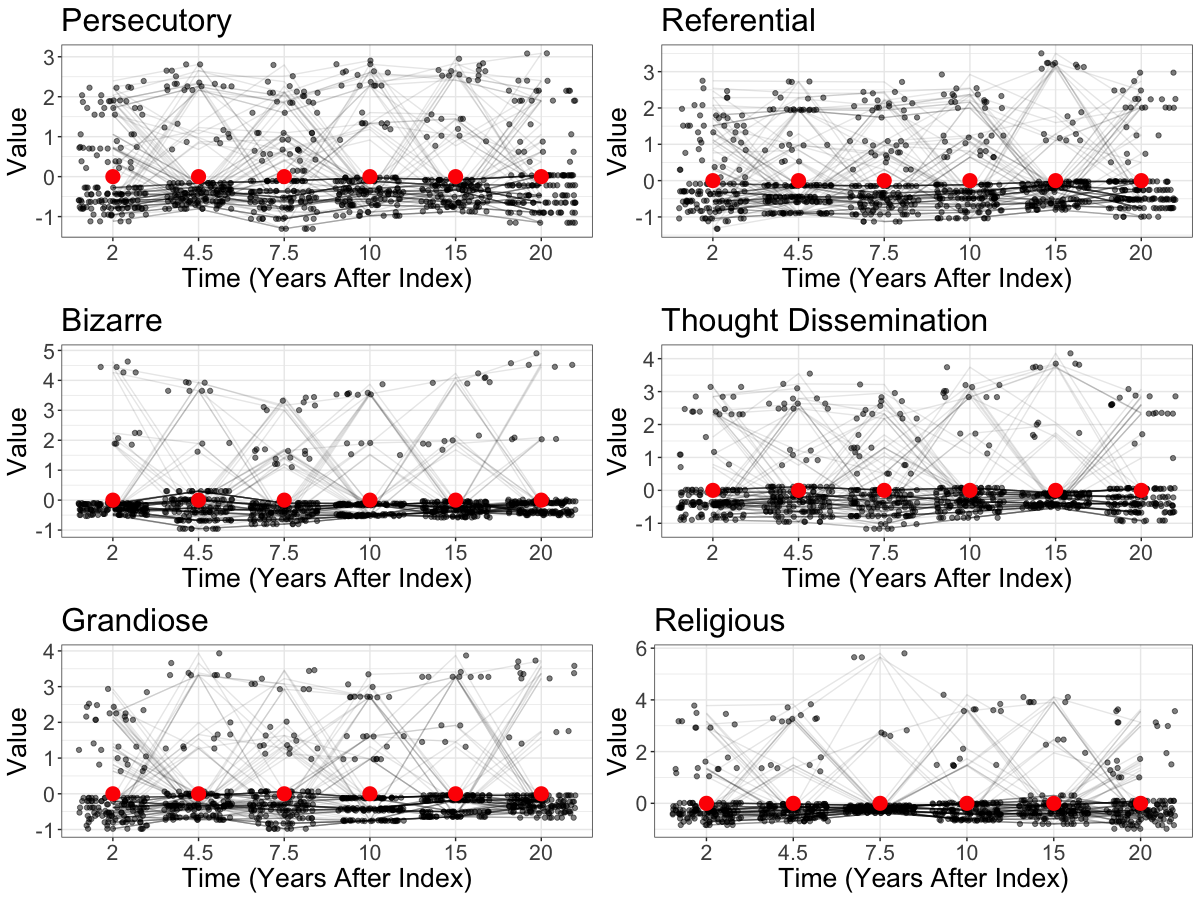

**Figure S2. Detrended and normalised between-individual residuals over time.**

Black dots and lines are individual participant scores for each time point. Red dots are grand averages across participants for each time point. Values are jittered for visual clarity and therefore some lines from timepoint to timepoint will be unlinked to dots within each time point on the x-axis. Participants were rated by a clinician as to whether delusions were not present, were borderline, or were clearly present. Plots demonstrate no linear trend in mean values across all time points. This data is controlled for by age, sex, IQ, or medication status.

**Figure S3. Delusions-only network model including nihilistic delusions.**

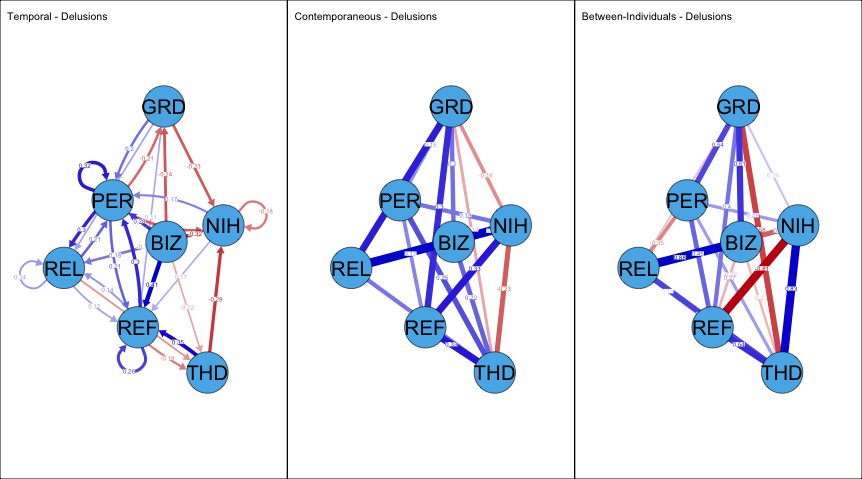

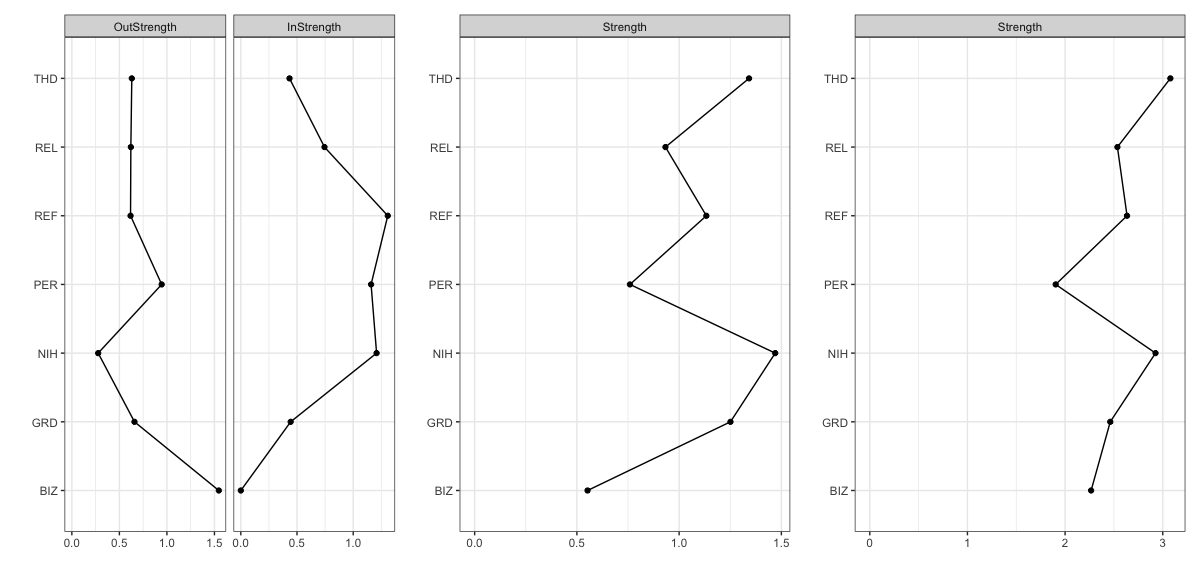

(A) A saturated network was approximated from the full dataset and had poor-moderate fit (RMSEA ~ 0.077, 95%CI [0.067, 0.080]; χ^2^(833) = 1442.22, p ~ 0; CFI = 0.88; TLI = 0.88). Thought dissemination (r = -0.29), persecutory (r = -0.22), bizarre (r = -0.32) and grandiose (r = -0.22) delusions all had significant (ps < 0.05) negative temporal relationships with nihilistic delusions. Nihilistic delusions were significantly negatively autoregressive (r = -0.18). Nihilistic delusions shared significant (ps < 0.05), positive contemporaneous associations with persecutory (r = 0.19), religious (r = 0.42) and referential (r = 0.33) delusions and shared a significant negative relationship with thought dissemination (r = -0.23). No significant between-subject relationship existed with nihilistic delusions. (B) Centrality estimates (absolute values) of each network.

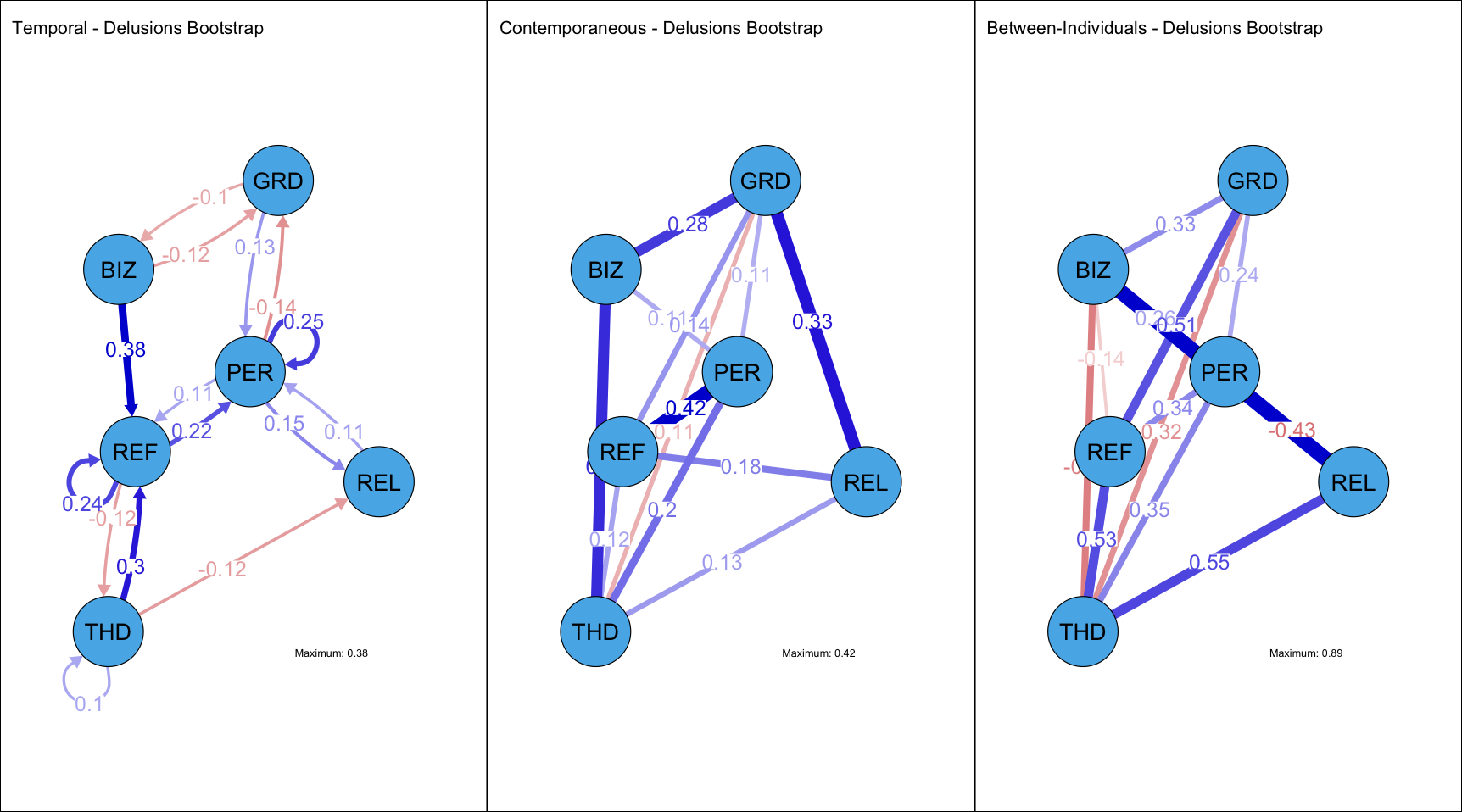

**Figure S4. Bootstrapped delusions-only network model.**

Over 500 iterations, 25% of the sample was randomly held out and the full model refitted on the remaining 75% of participants. Within each iteration the selected data imputed, each variable regressed against confounders, and scaled in the same manner as in the full model to control for errors and variance within the data cleaning and scaling process. The averaged edge weights and 95% confidence intervals (CI) over all 500 iterations were retained and reported. All edges that had 95%CI crossing zero were forced to 0.

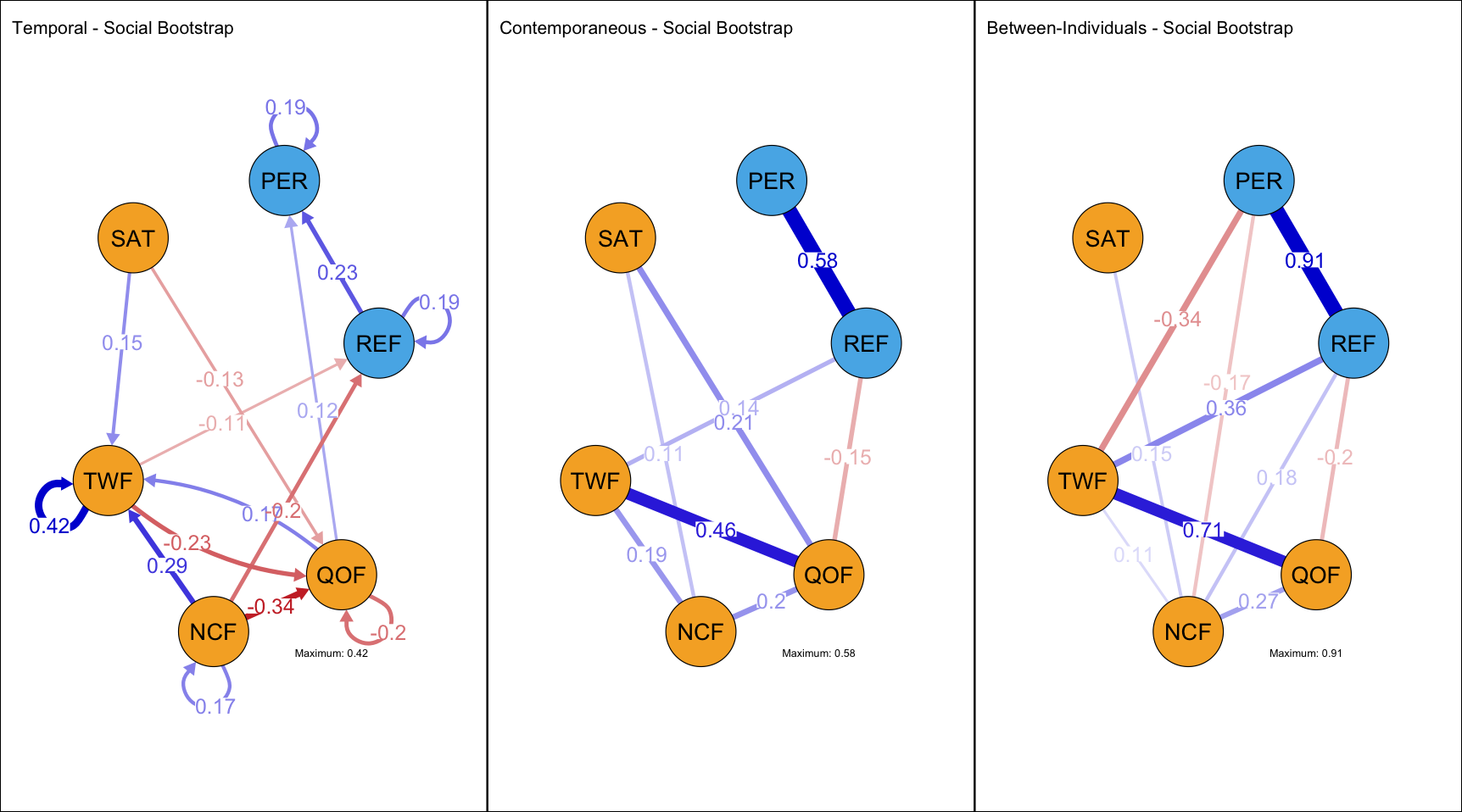

**Figure S5. Bootstrapped social variable network model.**

Over 500 iterations, 25% of the sample was randomly held out and the full model refitted on the remaining 75% of participants. Within each iteration the selected data imputed, each variable regressed against confounders, and scaled in the same manner as in the full model to control for errors and variance within the data cleaning and scaling process. The averaged edge weights and 95% confidence intervals (CI) over all 500 iterations were retained and reported. All edges that had 95%CI crossing zero were forced to 0.

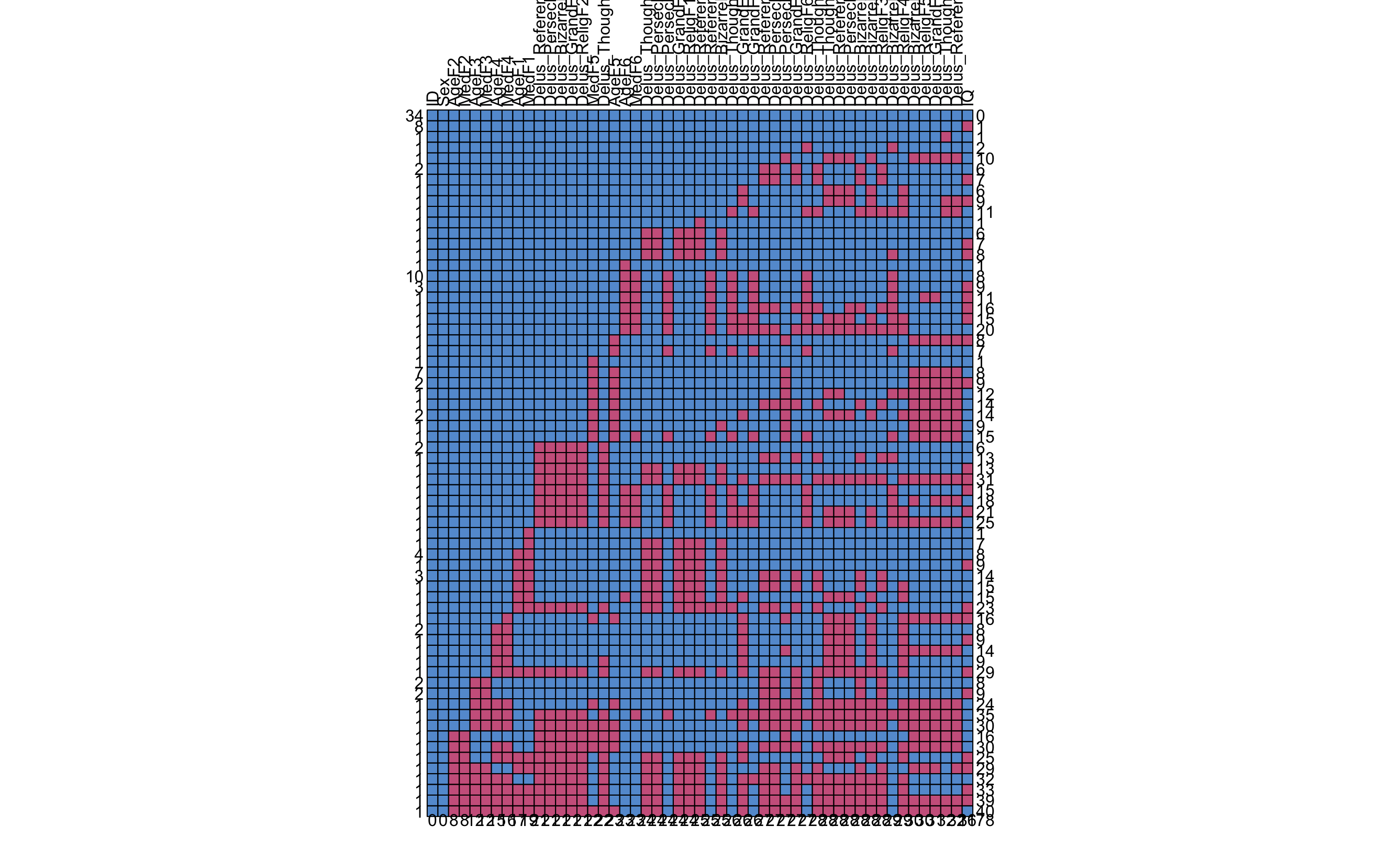

**Figure S6. Missing Data of The Delusions-Only Network.**

Metrics on the left-hand side on each row represent the number of participants for which the row is true for. The number on the right-hand side represent how many data points were missing. Columns represent variables. Numbers on the bottom represent number of missing data within columns. In total, out of 6885 data points across 135 participants, 17% (1178 points) were missing.

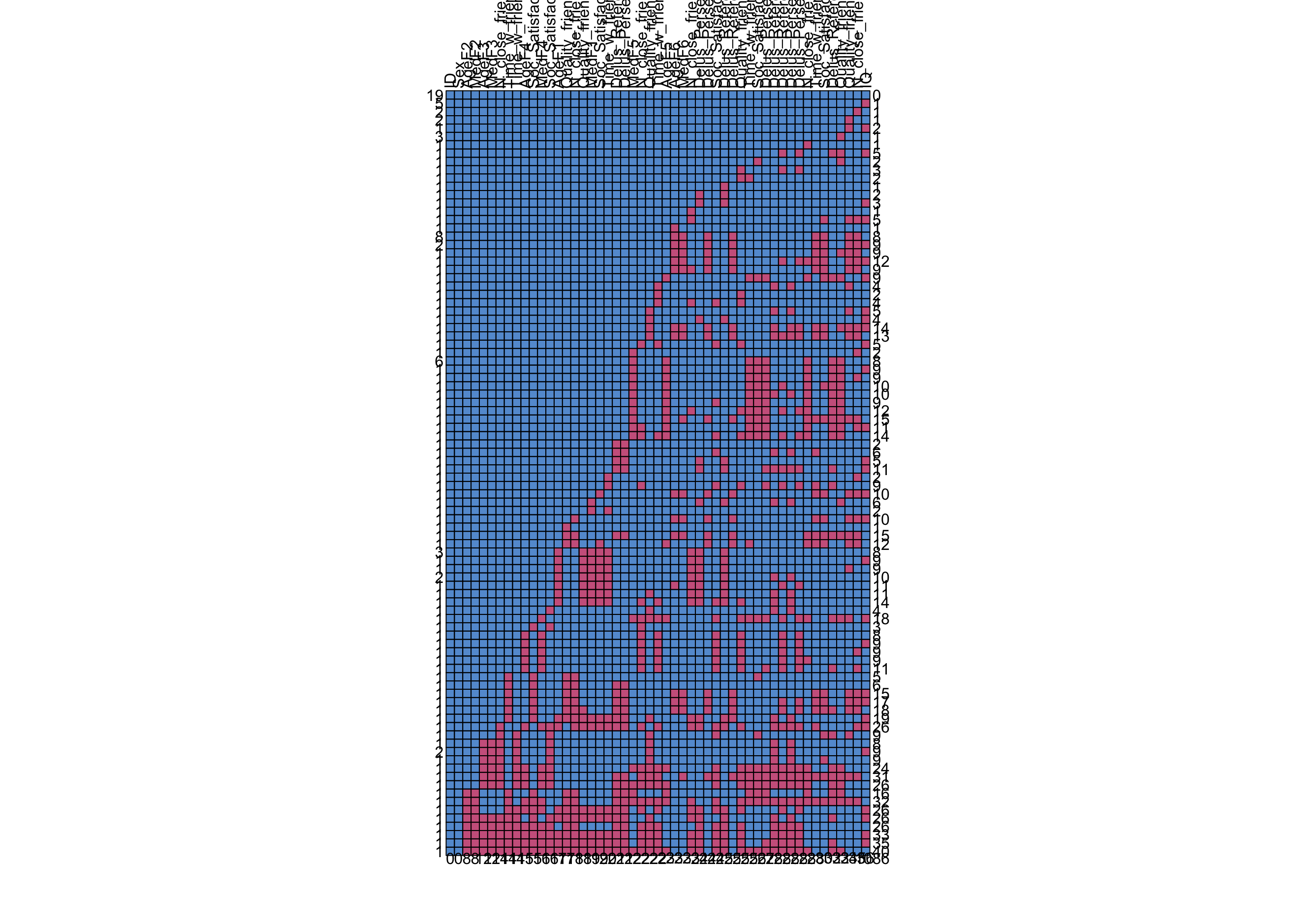

**Figure S7. Missing Data of The Social Network.**

Metrics on the left-hand side on each row represent the number of participants for which the row is true for. The number on the right-hand side represent how many data points were missing. Columns represent variables. Numbers on the bottom represent number of missing data within columns. In total, out of 6885 data points across 135 participants, 16% (1086 points) were missing.

**Table S1. Absolute bootstrapped centrality estimates [z scores].**

| **Delusions-Only Network** | | | | | | |
| --- | --- | --- | --- | --- | --- | --- |
| **1** | **2** | **3** | **4** | **5** | **6** |  |
| 0.74 | 1.24 | 0.76 | 0.96 | 0.68 | 0.93 | Between-Individual |
| 0.65 | 0.92 | 0.83 | 0.75 | 0.75 | 0.64 | Contemporaneous |
| 0.17 | 0.93 | 0.45 | 0.11 | 0.24 | 0.02 | Temporal-IN |
| 0.18 | 0.20 | 0.12 | 0.20 | 0.11 | 0.08 | Temporal-OUT |
| **Social Network** | | | | | | |
| 1.31 | 0.46 | 0.53 | 0.93 | 0.83 | 0.30 | Between-Individual |
| 0.57 | 0.48 | 0.48 | 0.72 | 0.73 | 0.39 | Contemporaneous |
| 0.32 | 0.35 | 0.09 | 0.70 | 0.68 | 0.02 | Temporal-IN |
| 0.26 | 0.04 | 0.24 | 0.41 | 0.28 | 0.07 | Temporal-OUT |
